# RT-HaND-C: A Multi-Source, Validated Real-World Head and Neck Cancer Dataset for Research

**DOI:** 10.1101/2025.05.08.25327208

**Authors:** Tom Young, Haleema Drake, Victoria Butterworth, Wulaningsih Wahyu, Bill Dann, Aga Giemza, Eleanor Ivy, Delali Adjogatse, Khrishanthne Sambasivan, Imran Petkar, Miguel Reis Ferreira, Anthony Kong, Mary Lei, Lisette Collins, Andrew King, Dika Vilic, Teresa Guerrero Urbano

## Abstract

**Background:** Real-world data (RWD) is essential in head and neck cancer (HNC) research, offering insights into outcomes among diverse, comorbid patients often underrepresented in clinical trials.

**Methods:** We developed RT-HaND-C, a multi-source clinical dataset integrating structured EHR data, unstructured data extracted using an AI-driven NLP tool, and previously manually-curated datasets, with extensive demographic, disease, laboratory, treatment, outcome and radiotherapy dosimetry data for all HNC oncology patients seen at our centre (2010–2023). The dataset underwent rigorous evaluation for accuracy, completeness, consistency, and usability.

**Results:** The retrospective cohort comprises 2,895 HNC patients with over 1.8 million data points across over 2000 data categories. Accuracy assessments exceeded 98% for most variables. An example of usability testing showing rapid extraction and evaluation of longitudinal weight patterns post-radical radiotherapy is depicted.

**Conclusions:** RT-HaND-C represents a novel, high-quality RWD resource and evaluation framework. The dataset is available for research and collaboration, with ongoing efforts to enhance completeness and support prospective updates.

## Introduction

The Food and Drug Administration (FDA) defines Real-World Data (RWD) as “data on patient health status and/or delivery of health care routinely collected from multiple sources outside typical clinical research settings” (1) such as Electronic Health Records (EHRs). RWD can be analysed to generate Real-World Evidence (RWE), increasingly applied for cancer research (2, 3). This drive is underpinned by several factors including the limitations of clinical trials, the gold standard practice-setting cancer research. Clinical trials provide vital insight into an intervention’s effect on patient outcomes but lack inclusivity, and therefore potentially generalisability, to increasingly elderly and co-morbid patient populations (4). Furthermore, trials are increasingly expensive to perform and may take many years to answer research questions (5). RWE may complement clinical trial research to address these issues. RWE generation is particularly relevant to head and neck cancer (HNC). HNC patients are disproportionately from lower socioeconomic backgrounds (6), and therefore may be particularly underrepresented in clinical trials (4). HNC patients can suffer significant morbidity post-treatment, with socioeconomic factors influencing long-term outcomes and survivorship. Consequentially, robust RWD offers an opportunity to capture a broader range of HNC patient experiences, directing future interventions to improve patient care.

Curation of robust RWD has challenges. RWD are collected during routine clinical practice and can therefore be inconsistent, incomplete (particularly compared to trial data) and subject to random and non-random recording errors and biases (7). RWD fragmentation, with patient data often held across multiple EHR systems, necessitates development of interoperable data pipelines. Furthermore, large proportions of important RWD exist in unstructured format within EHRs (8), such as text (e.g. “tumour showed lymphovascular invasion”) within a histopathology report. While EHR structured data may be rapidly and automatically extracted (9), gold standard for extracting unstructured RWD is through human abstraction (manual curation) but is unfeasible for large datasets due to time and therefore resources required to undertake at scale (2). Manual curation may also be error-prone (10).

Recent advances help address these issues. Initiatives to improve oncology data quality recorded in EHRs, such as Minimal Common Oncology Data Elements (mCODE), may facilitate data terminology and recording consistency across different institutions (11,12). Advancements in IT systems’ integration, including widespread adoption of EHR systems (13), have increased availability and accessibility of RWD sources, giving rise to the potential for rapid and automatic RWD extraction. Furthermore, Artificial Intelligence (AI) tools using Natural Language Processing (NLP) have been developed to curate unstructured data rapidly and accurately (14), albeit with limited application within cancer research to-date (15).

Many existing RWD sources are available. Large national datasets include the USA’s Surveillance, Epidemiology and End Results (SEER) Programme, which consists of 18 registries collecting cancer (including HNC) data for 48% of the US population (16). Large national-level datasets may however lack specific granular data available at the local institutional level. They also lack linked imaging and RT treatment datasets, of importance to multi-modal HNC research leveraging AI research techniques. Several HNC-specific real-world datasets (mostly unicentric) have been curated including the largest RADCURE (n=3346) (17), containing clinical and/or imaging data. These valuable resources do however generally contain only limited clinical data, which may limit their applicability to the many different research questions of importance to HNC patients. Further additional time-consuming data curation processes may therefore be required for future research projects.

Most current HNC datasets have reported limited clinical data validation. As per regulatory body data standards guidance (18, 19), robust validation of data quality is required before downstream RWE generation with valid conclusions. RWD quality dimensions required including dataset validity, completeness, uniqueness, accuracy, consistency and timeliness, which have been subcategorised into “relevance” and “reliability” criteria (2) (Figure 1). Adherence to FAIR principles, whereby scientific data is “Findable, Accessible, Interoperable, and Reusable” is key (20). ‘Live’ datasets, with updates incorporating additional data and prospective patients as time progresses, are of particular long-term value.

**Figure 1.**
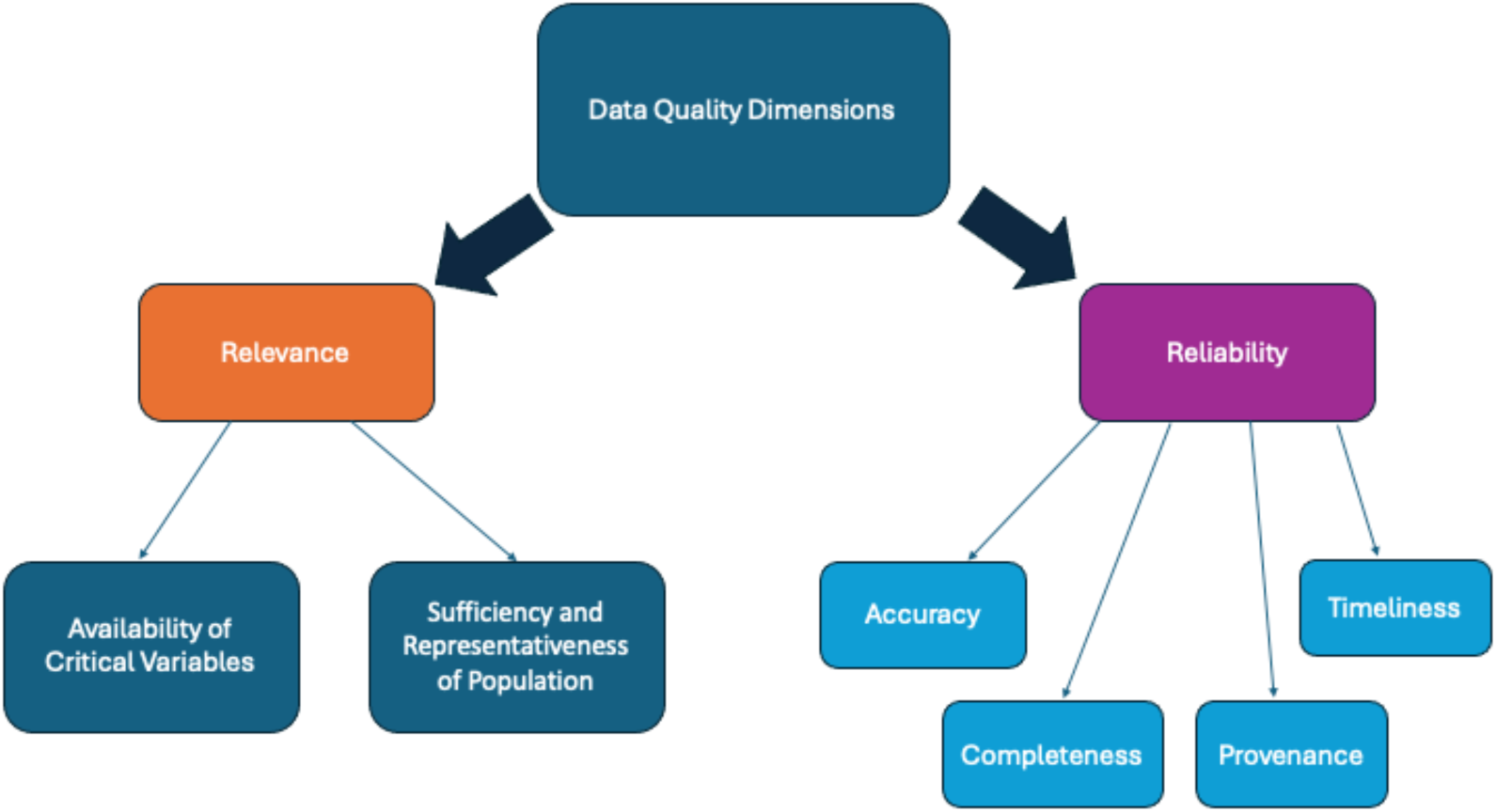
RWD quality dimensions as categorised by Castellanos et al. (2).

We aimed to address issues affecting currently available datasets by leveraging the vast amount of data available from our HNC oncology patient cohort at our centre, serving a population of 2 million, to develop RT-HaND-C (RadioTherapy – Head and Neck Dataset – Clinical): a dataset containing clinical data for our entire oncology HNC cohort seen since 2010, containing demographic, disease, treatment, outcome, laboratory and RT dosimetry. We then implemented a rigorous validation, verification and usability evaluation process to ensure data quality for future research use. We linked RT-HaND-C with its corresponding imaging dataset (RT-HaND_I: Radiotherapy – Head and Neck Dataset – Imaging (21)) containing both diagnostic and RT images to facilitate multi-modal research projects for internal and external researchers. Finally, we aimed to ensure our ‘live’ dataset incorporated an automatic clinical data curation pipeline from EHR systems for prospective data.

## Materials and Methods

### Population

Inclusion and exclusion criteria were applied to comprehensively capture the HNC oncology patient population, consisting of patients referred to the clinical oncology service for consideration of radiotherapy or systemic anti-cancer therapy (SACT), for the initial dataset cohort (Figure 2). The date range for this initial dataset cohort was 1/1/2010 to 4/10/2023. A record (‘HNC Masterlist’) of all patients seen by the HNC oncology team since 2010 was used to establish the cohort to then apply inclusion/exclusion criteria. This record had been updated twice weekly since its creation with relevant HNC patient information (including identifying and limited clinical information). Cohort completeness was ensured by cross-referencing against other sources, for example structured data extracts for all patients undergoing HNC RT from the RT system.

**Figure 2.**
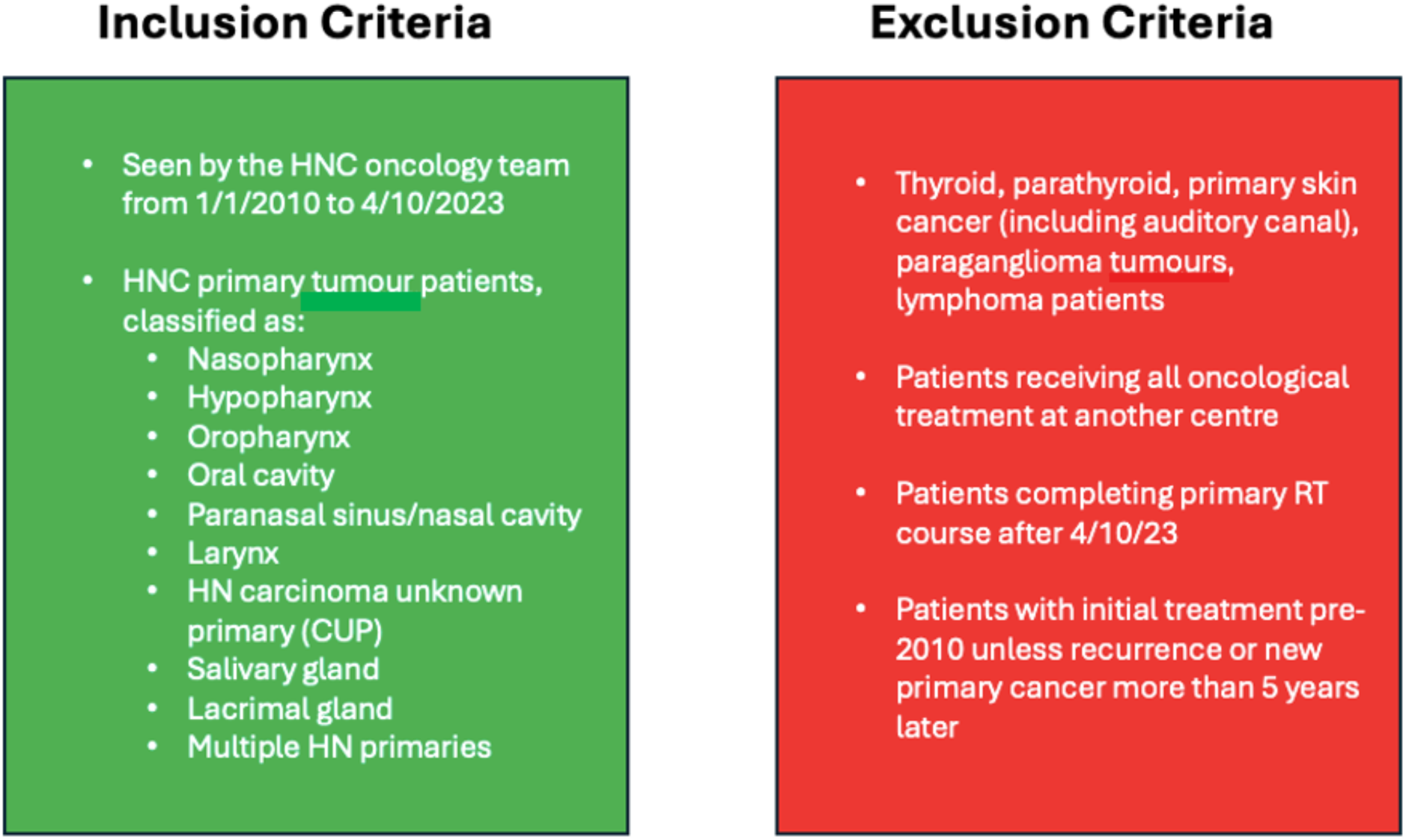
Inclusion and exclusion criteria applied for identification of initial cohort of interest. Patients after 4/10/23 subsequently added to cohort as dataset updated prospectively.

The end date was set as a new unifying EHR system (Epic (22)) was deployed on 5/10/23 to supersede multiple legacy EHR systems. It was therefore imperative to build a clinical dataset for our patients treated prior to Epic go-live whilst legacy EHR data remained easily accessible. Once Epic data warehousing solutions were fully integrated this dataset could then be linked with data pipelines to both update the existing cohort with prospectively collected data (such as outcome data) and add new HNC oncology patients.

### Data Categories

With reference to mCODE^TM^ Version 4 (to ensure interoperability and consistency) required data categories were identified (Figure 3). The full list of data categories can be found on Github, with this data dictionary ensuring data consistency and standardisation (https://github.com/GSTT-Radiotherapy-Physics/RT-HaND/blob/main/Documentation/Appendix/07a%20RT-HaND_C%20Retrospective%20Data%20Dictionary.xlsx).

**Figure 3.**
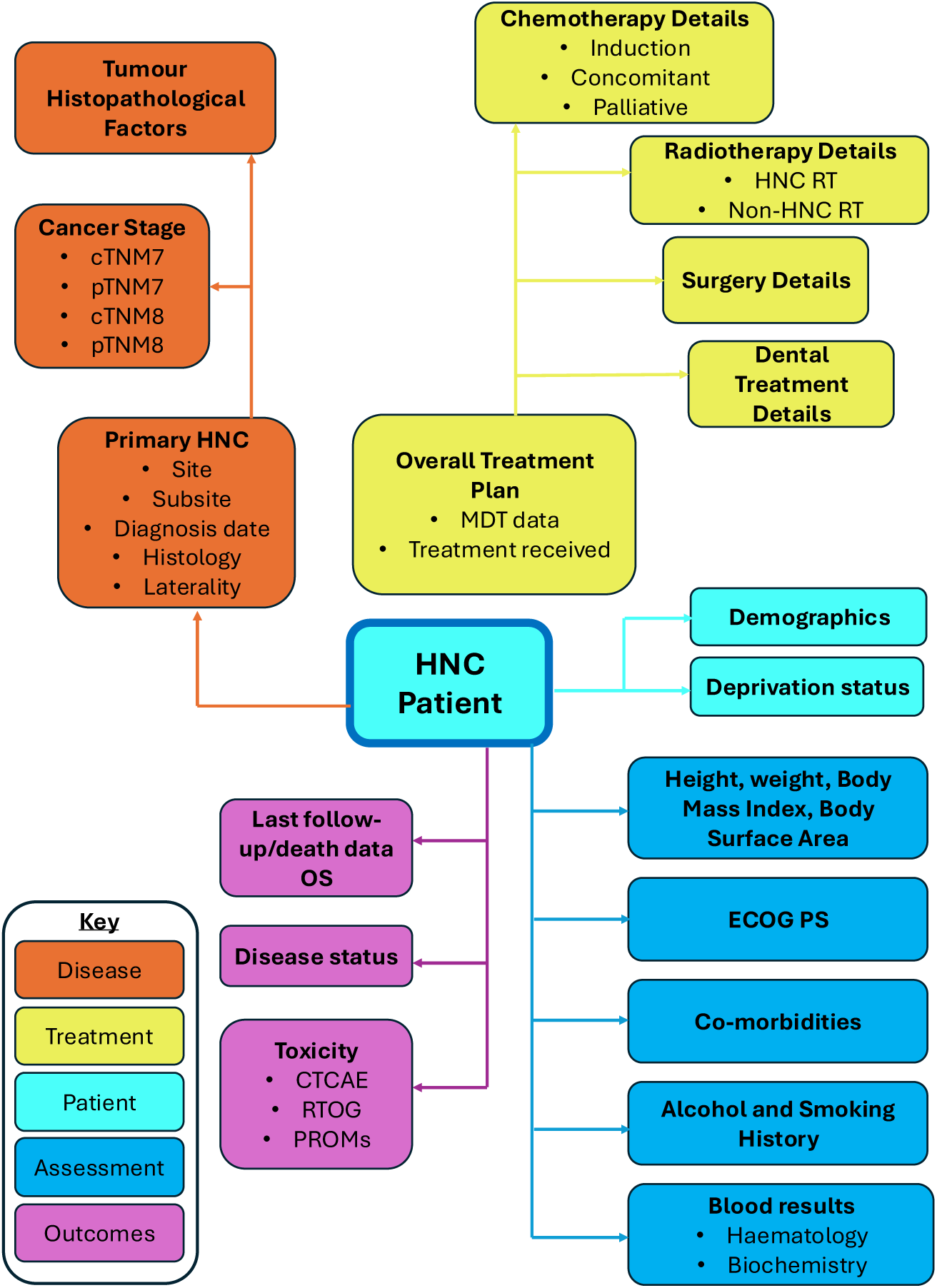
Summary of data present in retrospective HNC clinical dataset (corresponding to mCODE data categories (11).

### Data Curation Sources

Where available, methods utilising automatic data curation at scale were favoured. RT-HaND-C was populated from multiple data sources (Figure 4).

**Figure 4.**
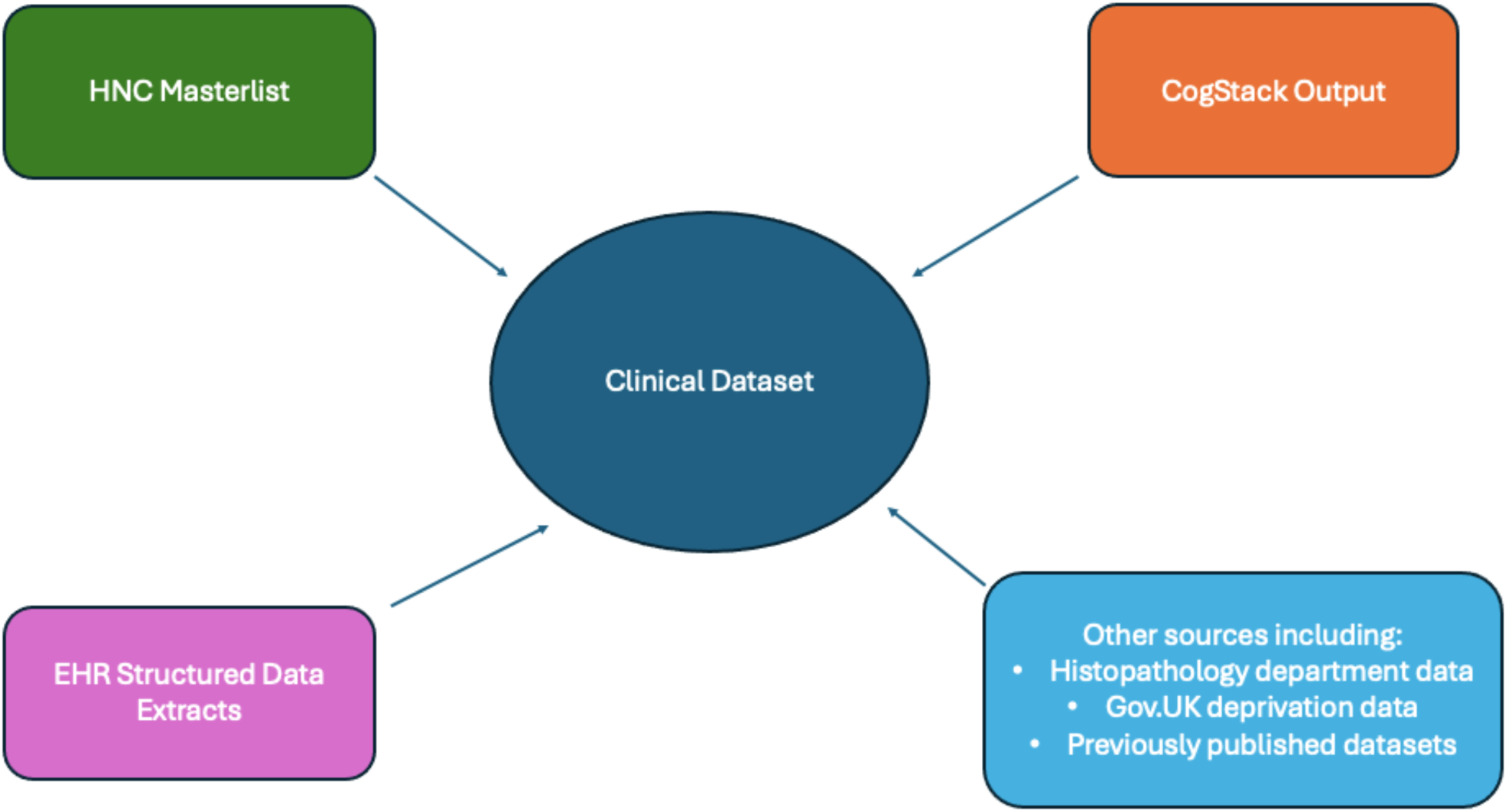
Data sources used for the retrospective clinical dataset.

#### i) Structured Data Extraction from EHRs

HealthCatalyst, a platform utilised to hold clinical data tables from different legacy EHR systems on a single Electronic Data Warehouse (EDW) (9), was used to rapidly extract relevant structured data required for clinical dataset population with Structured Query Language (SQL). All legacy EHR system EDW data tables of interest were up-to-date to 4/10/23 in accordance with the initial cohort’s required final assessment timepoint. EHR systems containing data extracted in this method were MOSAIQ® (cancer patient EHR), iSOFT Clinical Manager (test result and clinical document EHR), and Personal Information Management System (patient encounter EHR). An “Extract, Transform, Load” (ETL) data curation process (23) was followed to format data as required. Large amounts of demographic, disease, treatment (dental, surgery, chemotherapy, radiotherapy), blood result, RT dosimetry and outcome (death, follow-up, toxicity assessments completed at point-of-care) data were rapidly curated in this way. Postcode data was linked to deprivation indices data for each patient using the UK Government lookup tool (24).

#### ii) Unstructured Data Extraction using NLP

Cogstack, a previously evaluated AI tool by our group (25), was used to extract HNC-specific concepts from unstructured clinical notes/letters/reports for HNC patients. CogStack was run for co-morbidity, disease, treatment and outcome concepts with validated performance for our patient population across relevant clinical documents within EHR systems to provide an output of the number of concept pick-ups within the specified searched EHR system documents for each patient. This was then converted to a binary output of “0” (concept not present) or “1” (concept present) (using a pre-set thresholding strategy), and integrated into RT-HaND-C. The full list of SNOMED CT concepts used can be seen in Supplementary Table 1.

#### iii) Manually Curated Data

Limited data was available from previously manually curated datasets, which were therefore integrated into RT-HaND-C. This data was particularly useful for categories not curatable using approaches i) and ii). For example, for post-operative HNC patients’ surgical margin was not stored in structured form, nor adequately extracted by CogStack. Several previous internal projects had retrospectively curated disease and outcome data. These datasets included 172 oral cavity cancer patients (26), 69 larynx cancer patients (unpublished data), 524 oropharynx cancer patients (27) and PD-L1 testing data (unpublished data). This valuable source of previously manually curated data was therefore incorporated into RT-HaND-C.

### Data Integration

All relevant clinical data was integrated together using HealthCatalyst’s SAM Designer tool (9), forming a complete initial cohort dataset from which required data could be extracted as a CSV file for analytic projects. RT-HaND-C could be searched by applying filters (for example, selecting only HPV-positive oropharynx cancer patients) tailored to data required for specific research projects. RT-HaND-C was also virtually linked to a large imaging dataset (RT-HaND-I) containing diagnostic and RT imaging data (Figure 5), described in detail elsewhere (21) and stored on the Extensible Neuroimaging Archive Toolkit (XNAT) platform virtually linked using common primary identifiers.

**Figure 5.**
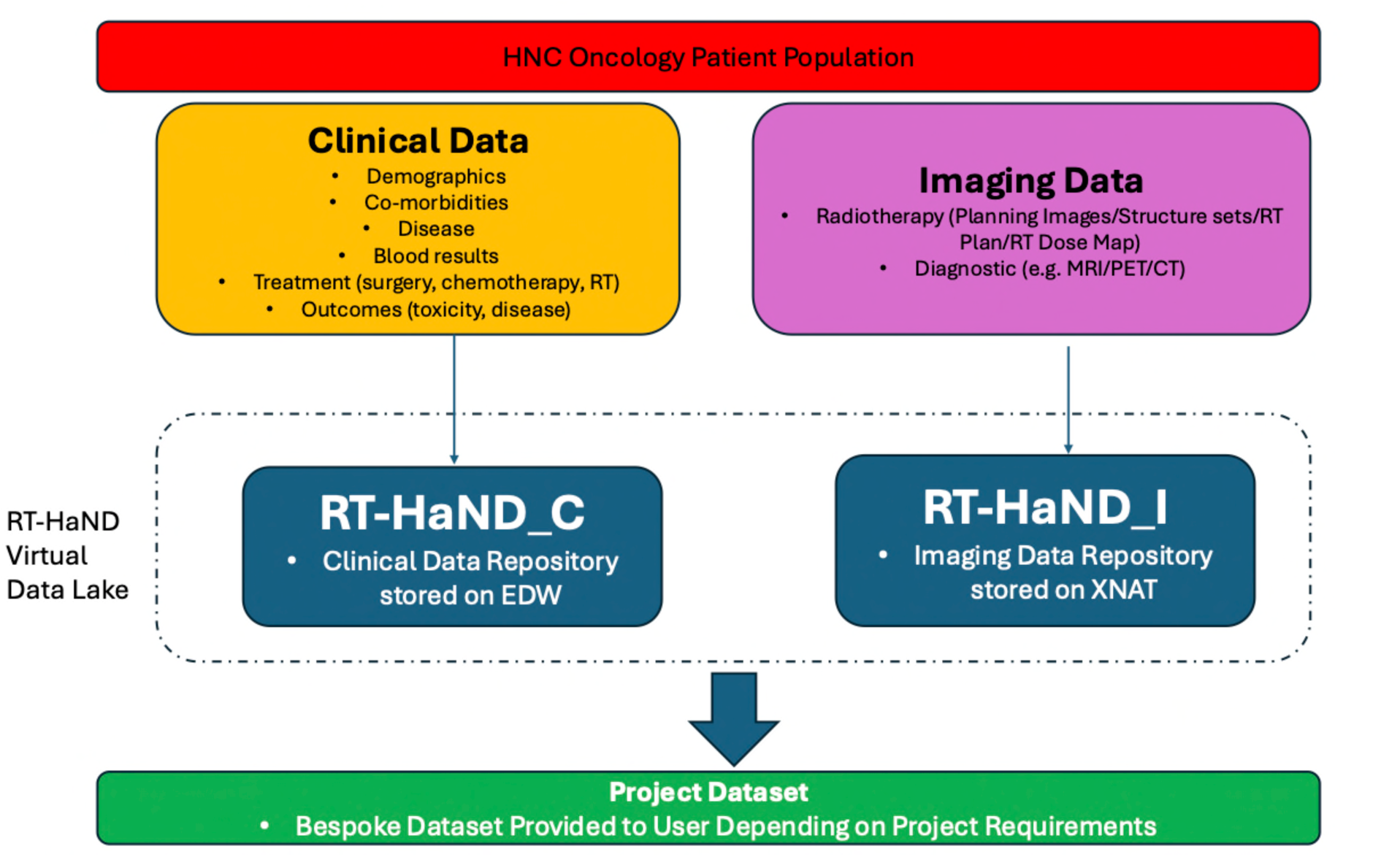
Workflow for HNC RT patient clinical and imaging datasets.

### Dataset Evaluation

RT-HaND-C was assessed for dataset accuracy, timeliness, completeness, consistency and usability. As per published recommendations (2), this process was performed both throughout the dataset build and as a final validation assessment. Following data validation, any incorrect data found were subsequently corrected within the dataset.

#### i) Data accuracy and timeliness

Random samples (n=60) were selected for each data category. For each patient randomly selected, the data category being assessed was manually corroborated against the EHR, and the data point was recorded as correct or incorrect to calculate accuracy (and for binary categories, positive and negative predictive value (PPV/NPV)). For timeliness, the timepoint of relevance taken for assessment against was 4/10/2023. Only complete data within this 60 patient sample was assessed for accuracy – for example, if 20 cases were missing data, accuracy was assessed and calculated for the remaining 40 patients.

#### ii) Data completeness assessment

Percentage of complete and incomplete data points was calculated for each data category, taking into consideration that incomplete data did not necessarily mean missing data; for example, a missing TNM staging would likely represent truly missing data (all cancer patients should undergo TNM staging), whereas absent 1-year post-treatment assessment data may have never been possible if a patient had died before 1-year.

#### iii) Data consistency assessment

Consistency of data was assessed against data dictionary definitions to ensure values recorded were denoted in a standardised manner for each data category.

#### iv) Usability assessment

As a use-case assessment, we aimed to use RT-HaND-C to answer a clinical question. There is currently limited data in the literature on long-term effects of primary IMRT on weight, beyond the acute setting immediately post-RT. We therefore assessed RT-HaND-C’s usability to identify a clear population (primary IMRT HNC patients) and provide relevant data for these patients (weight at baseline, and different timepoints during/after RT), for further analysis to address the research question. Longitudinal weight values were assessed for the cohort. Wilcoxon Signed-Rank Test was used to compare paired weights at baseline and other timepoints. Spearman’s correlation was used to assess correlation between maximum acute period weight loss from baseline compared to later timepoints post-RT.

### Permissions

Permission to undertake this work was approved by our centre’s Information Governance team following completion of a Data Protection Information Assessment. Patient data was used as per the opt-out consent process underpinned by our centre’s Research Ethics Committee ethics approval process (REC Reference:18/NW/0297, IRAS Project ID: 231443) (28).

## Results

### Dataset Overview

RT-HaND-C consisted of over 1.8 million data points for the initial cohort of 2895 patients seen by the HNC oncology team meeting inclusion/exclusion criteria across 2114 data categories.

Figure 6 shows an overview of the population characteristics (tabular data displayed in Supplementary Table 2). The majority of the cohort’s HNC patients are male, with oropharynx cancer being the most common tumour site. New patients per year gradually increased over the set time-period, with a notable dip seen in 2020 coinciding with the Covid-19 pandemic. Figure 7 demonstrates 5-year overall survival for the entire patient cohort.

**Figure 6.**
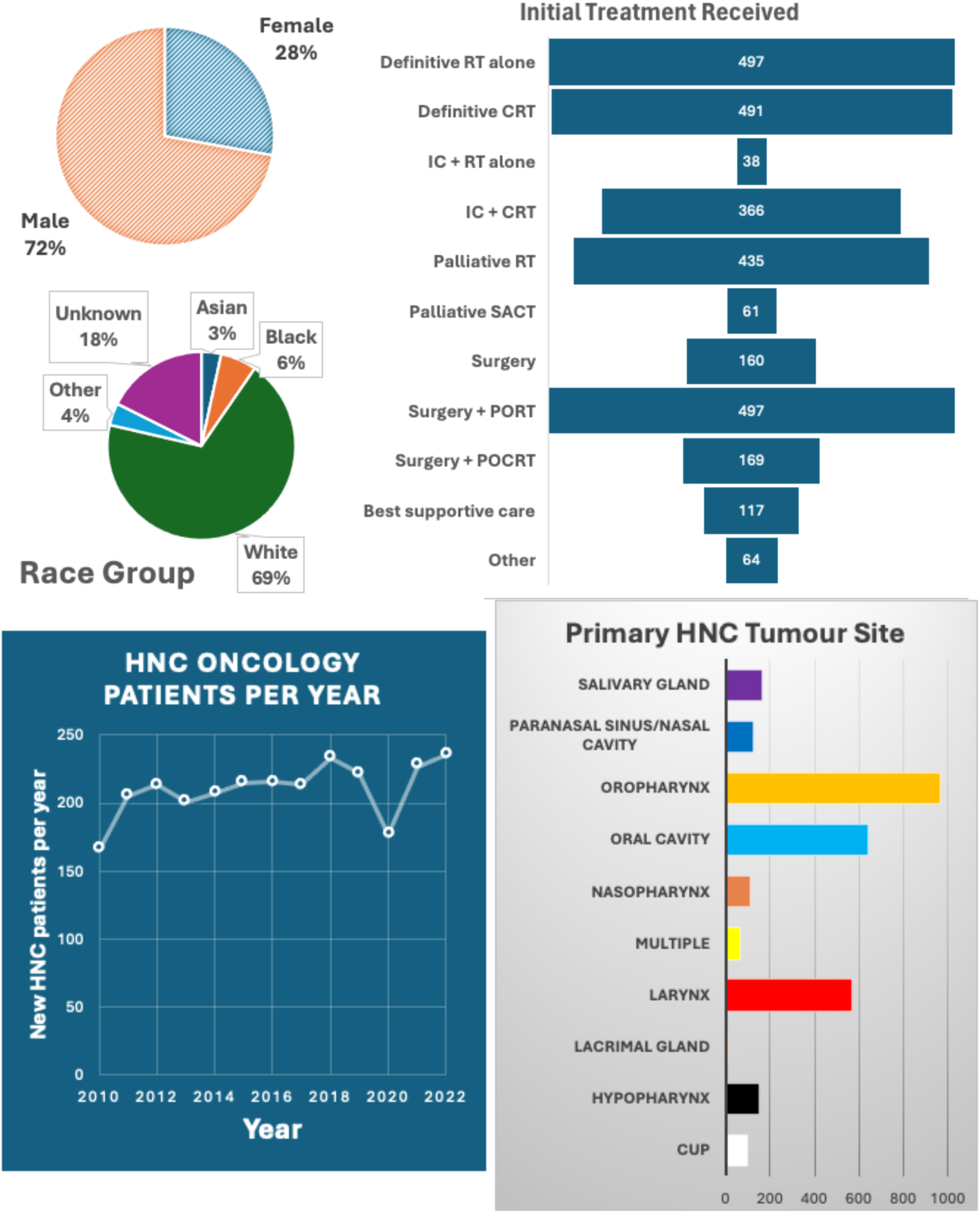
Infographic demonstrating the Head and Neck Oncology Patient Population comprising RT-HaND-C’s population characteristics. Treatment abbreviations: CRT = chemoradiotherapy, IC = induction chemotherapy, SACT = systemic anti-cancer therapy, PORT = post-operative RT, POCRT = post-operative chemoradiotherapy. Disease abbreviations: CUP = (Head and Neck) Cancer of Unknown Primary.

**Figure 7a.**
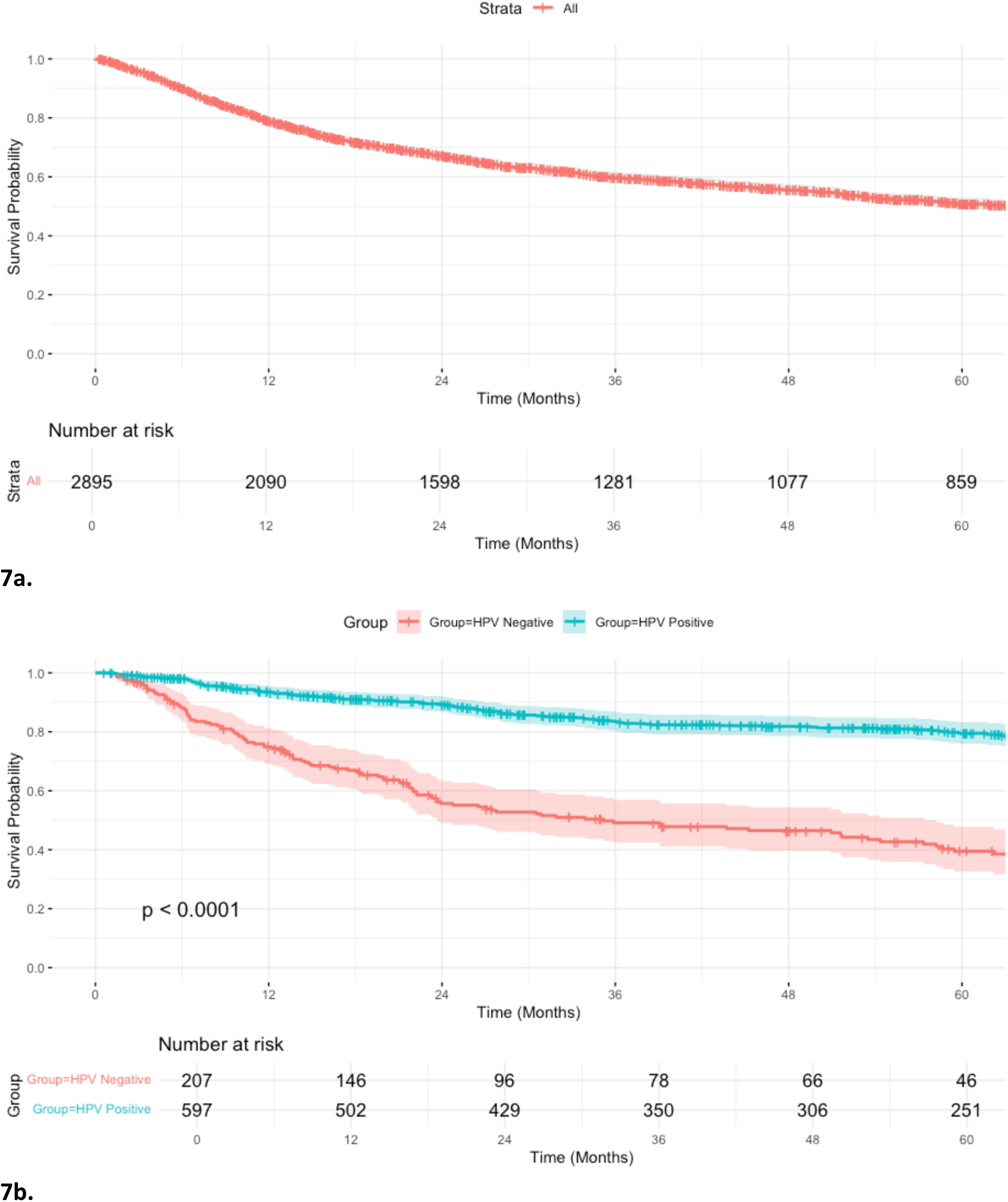
Kaplan-Meier curve demonstrating 5-year overall survival for RT-HaND-C’s entire initial patient cohort. **7b.** Example subgroup analysis: Kaplan-Meier curve demonstrating 5-year overall survival for RT-HaND-C’s initial cohort of all HPV-positive and HPV-negative oropharynx cancer patients. Log rank test used to compare survival curves.

### Evaluation

#### i) Data accuracy and timeliness

Overall assessed data accuracy was ≥98% (Figure 8, Supplementary Tables 3-9 for full accuracy assessment results). Data for 137 of 193 data categories assessed was 100% accurate. PPV and NPV was >0.95 (median PPV and NPV of 1) across all binary categories assessed. Co-morbidity data had the lowest overall accuracy (98%). Assessed demographic data was 100% accurate apart from postcode (1 patient moved, but change was not recorded in all EHR systems).

**Figure 8.**
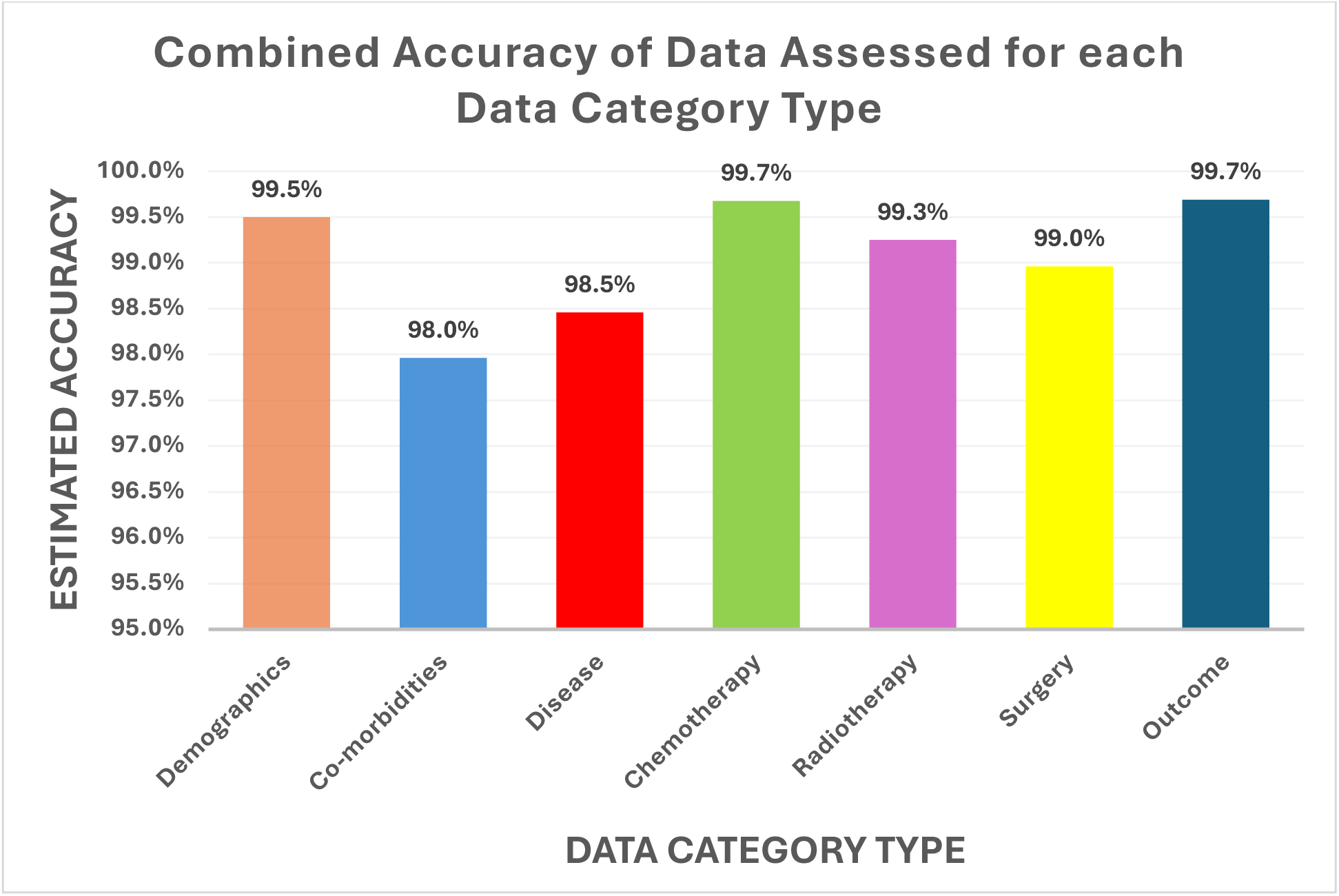
Summary of estimated accuracy of each data category based on data accuracy assessment of complete data within dataset.

Disease characteristic data was 95% accurate for all subcategories assessed except disease laterality (93.1%) and perineural invasion (85%). Overall treatment received, chemotherapy, RT, surgical and outcome data was estimated to be > 95% accurate for all categories. For data directly extracted without transformation (e.g. patient height values), all data assessed reflected exact EHR source data, with no data alterations/corruption encountered. All data was up-to-date as of 4/10/23.

#### ii) Data completeness

Key categories such as diagnosis and date, disease site, histopathology, initial treatment received, treatment data (surgery, RT and chemotherapy), and follow-up (last follow-up, death status/date) data was 100% complete. Completeness varied according to data availability across different data categories (available at: https://github.com/GSTT-Radiotherapy-Physics/RT-HaND/blob/main/Documentation/Reference%20Docs/15e%20RT-HaND_C%20Retrospective%20Dataset%20Build%20SOP%20Appendix%205-%20Completion%20Statistics.docx). Data completeness was low (<10%) for a small amount of data categories where NLP performance was inadequate and no structured data was available, such as specific histopathological features including lymphovascular invasion.

#### iii) Data consistency

Terminology within the clinical dataset was consistent across all data categories according to data dictionary definitions set pre-build. This included absent data to denote whether this data was missing (‘NULL’) or never existed (‘Not applicable’).

#### iv) Usability

RT-HaND-C’s finalised initial cohort dataset was found to be rapidly searchable (<2 minutes) to identify the defined subgroup of interest (radical VMAT/IMRT patients, n=1154) and extract required data (including 6750 weight values at different timepoints) for this subgroup for further analysis purposes. Baseline (pre-RT) weight values were present for 1152 of 1154 patients. Shapiro-Wilk Test found weight values demonstrated significant deviation from normality (positively skewed) for all timepoints except 5-years post-RT. Median baseline weight was 75.7kg (Lower quartile (LQ) 64.0kg, upper quartile (UQ) 88.7kg). Paired weight values at different timepoints post-RT were compared to baseline weight: Peak weight loss was seen at 6-months post-RT (10.1% median reduction from baseline) (Table 1). Wilcoxon Signed-Rank Test demonstrated significant reduction in weight at all timepoints, gradually normalising but persisting up to 5-years (Figure 9).

**Table 1.** Weight changes post-RT: Median weight difference (kg and percentage) at different timepoints post-RT when compared to baseline weight, with lower quartile (LQ) and upper quartile (UQ). Statistical comparison with Wilcoxon Signed-Rank Test, significance value with Bonferroni correction applied p<0.00417 (significant differences from baseline highlighted in green). Percentage of patients with more than 5% and 10% weight loss observed shown.

| Timepoint | Number of available comparisons against baseline weight | Median weight difference from baseline weight (kg (LQR-UQR)) | Median weight difference from baseline weight (% (LQR-UQR)) | Comparing with baseline weight: p-value | %pt >5% WL | %pt >10% WL |
| --- | --- | --- | --- | --- | --- | --- |
| Penultimate week RT | 811 | -3.1 (-5.4 to -0.8) | -4.1% (-7.0% to -1.2%) | <0.001 | 42.5% | 8.4% |
| Final week RT | 619 | -4.2 (-7.2 to -1.8) | -5.7% (-9.2% to -2.5%) | <0.001 | 54.8% | 19.5% |
| 1wk post-RT | 493 | -5.3 (-8.4 to -2.6) | -7.0% (-10.5% to -3.5%) | <0.001 | 65.9% | 28.0% |
| 2wk post-RT | 522 | -6.1 (-9.4 to -3.1) | -8.0% (-11.7% to -4.3%) | <0.001 | 72.0% | 36.0% |
| 6wk post-RT | 613 | -6.3 (-10.1 to -3.3) | -8.7% (-12.4% to -4.5%) | <0.001 | 71.9% | 41.3% |
| 3m post-RT | 653 | -7.2 (-11.6 to -3.0) | -9.5% (-14.4% to -4.4%) | <0.001 | 73.0% | 47.9% |
| 6m post-RT | 493 | -7.3 (-12.1 to -3.1) | -10.1% (-14.9% to -4.6%) | <0.001 | 74.0% | 50.5% |
| 1yr post-RT | 479 | -6.8 (-11.7 to -1.7) | -8.9% (-14.3% to -2.5%) | <0.001 | 65.8% | 46.3% |
| 2yr post-RT | 353 | -3.9 (-8.3 to 0.2) | -5.7% (-10.4% to 0.3%) | <0.001 | 53.5% | 28.3% |
| 3yr post-RT | 231 | -3.1 (-8.4 to 1.2) | -4.0% (-10.2% to 1.8%) | <0.001 | 46.3% | 26.0% |
| 4yr post-RT | 180 | -3.4 (-8.6 to 0.1) | -4.3% (-10.7% to 0.2%) | <0.001 | 47.8% | 26.7% |
| 5yr post-RT | 151 | -2.2 (-7.8 to 1.7) | -3.2% (-10.1% to 2.5%) | <0.001 | 46.4% | 25.2% |

**Figure 9.**
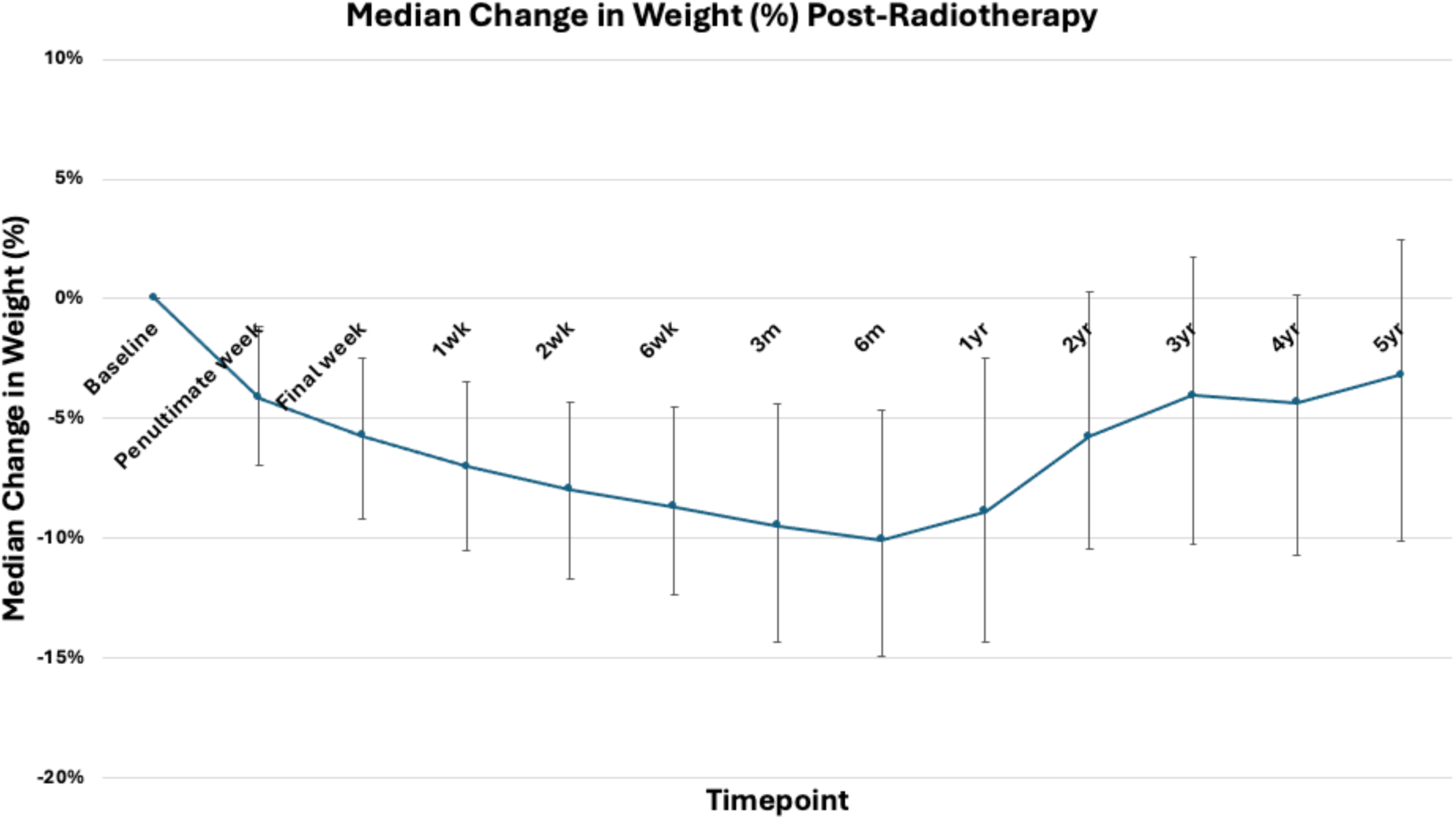
Median weight change (percentage of body weight) post-radiotherapy.

When considering the minimum weight value recorded from the ‘acute period’ (the penultimate or final week of RT, or 1-or 2-weeks post-RT), the median ‘acute period’ maximum weight loss was 5.45kg (7.4%). To analyse whether disease recurrence confounded for long-term weight loss trends observed, weight loss at 5-years was assessed excluding patients with recurrence within 5-years post-RT. 13 of 151 patients had recurrent disease (100% accurate on manual corroboration with EHR data) denoted by RT-HaND-C. Wilcoxon Signed-Rank test demonstrated significant weight loss (median loss 2.1kg (3.0%)) at 5-years even when excluding relapsed patients (p<0.001).

Correlation analysis was performed to assess the relationship between ‘acute period’ maximum percentage weight change with percentage weight change at later timepoints. This demonstrated a gradually reducing but significant correlation between the acute period weight change and all timepoints (except 4-years post-RT), including 5-years post-RT (Supplementary Table 10).

## Discussion

### Overview and Strengths

We built an extensive clinical dataset containing over 1.8 million demographic, co-morbidity, diagnosis, treatment, laboratory and outcome data points for a complete initial cohort of the 2895 HNC oncology patients. Evaluation demonstrated high accordance to data quality dimensions (1, 2, 29), with high accuracy, completeness and consistency demonstrated. RT-HaND-C is therefore a relevant longitudinal dataset representing a diverse real-world HNC oncology population.

RT-HaND-C has several strengths, including incorporation of data categories not present in other HNC RWD datasets (30, 31). Comprehensive subcategories of demographics (including deprivation), co-morbidities, diagnosis factors, treatments received, laboratory results, and outcomes, provide a wide-ranging patient-factor focused clinical dataset well-placed to facilitate robust RWE generation. This is therefore a scaled dataset with potential application to many different research questions. For example, RT-HaND-C contains over 300,000 data points for physician-reported toxicity grading data documented prospectively at the point-of-care. To our knowledge this represents one of the largest amalgamations of longitudinal HNC toxicity grading and represents just one example of the scaled data available. Virtual linkage of clinical data to diagnostic and RT imaging data (RT-HaND-I, containing 22,170 imaging studies for the initial cohort), facilitates data usage for multi-modal research projects. While other groups have developed multi-modal datasets of similar patient numbers, such as RADCURE (17), RT-HaND-C contains a broader range of clinical data categories.

We deployed several strategies during dataset build to ensure data robustness. We followed regulatory body RWD guidance to ensure data curation and evaluation was undertaken following gold standard approaches. An innovative approach in the dataset build was using an AI tool, CogStack, to convert unstructured data sources (e.g. clinical letters) into structured data for RT-HaND-C for previously validated SNOMED CT concepts. To our knowledge, this is the first UK oncology clinical dataset part-curated using NLP outside of a commercial setting. This is a method used by leading multi-billion dollar commercial companies such as Flatiron Health (2, 32), that we have managed to replicate despite having significantly more limited resources available to us. This approach demonstrates a working example that NLP can be used in conjunction with structured data extracts for dataset curation.

Cross-referencing of multiple data sources allowed targeted and feasible verification checks to maximise data quality. For example, chemotherapy data was principally from an EHR structured data extract but was also cross-checked with CogStack’s output for concepts such as ‘induction chemotherapy’ and ‘concomitant chemotherapy’. This process facilitated identification of missing data (for example those who had chemotherapy paper prescriptions prior to electronic prescribing in 2012 and therefore missing from EHR chemotherapy prescription data extracts).

Price et al. highlighted demonstrating RWD impact to motivate clinicians to engage with RWD research is critical to continued success, leading to both improved data quality at the point of capture and increased dataset usage for research (33). The usability assessment here demonstrated RT-HaND-C to be rapidly searchable to extract relevant data categories for a defined population of interest. We noted a gap in the existing literature regarding long-term weight trends post-RT (particularly in the IMRT era), with most relevant studies investigating small patient cohorts during and immediately post-RT. RT-HaND-C was able to address this as a demonstration of potential impact. Data analysed demonstrated novel findings; significant weight loss was seen at all timepoints persisting up to and including 5-years, even when excluding recurrent disease patients at this timepoint. Although median weight difference from baseline gradually normalised over time, data demonstrates that for many definitive IMRT patients weight loss experienced during HNC RT remains a long-term phenomenon. Our data also revealed that peak weight loss was seen at 6-months post-RT, considerably later than most acute toxicities affecting oral intake typically resolve by, suggesting potential importance of other factors such as low mood and anorexia. While explanatory work was beyond the focus of this dataset usability and demonstration work, findings underscore the chronic nature of weight loss after IMRT and warrant further exploration which our group will undertake using RT-HaND in the future.

### Limitations

RT-HaND-C currently has limitations, reflective of challenges associated with RWD. Firstly, RT-HaND-C is a unicentric dataset representing our centre’s HNC oncology population, but not necessarily other centres’. It contains only HNC patients seen by the oncology team for consideration of radiotherapy, SACT and/or best supportive care, rather than all our centre’s HNC patients. There are several areas where work is ongoing to improve RT-HaND-C, primarily relating to data not available within a structured data extract, nor extractable using NLP/CogStack. While the use of CogStack’s NLP ability was valuable, it was not without limitations. CogStack provided binary present/absent data for concepts, but did not directly provide temporality (for example date of a procedure or treatment). This limited granularity of data captured such as detailed disease outcome data beyond follow-up and death dates. For example, CogStack’s output demonstrated whether ‘disease recurrence’ had occurred for a patient, but not date of recurrence. Likewise, CogStack was not previously validated in extracting histopathological report concepts such as presence of lymphovascular invasion.

Some of these data categories were available from previously manually curated datasets however completeness assessment demonstrates this to be an area for ongoing improvement. Future NLP advancements may resolve this issue but targeted manual curation may be necessary to cover relevant gaps in the short-term.

Data curation challenges were seen for more historic patients (particularly pre-2012), where data completeness was observed during curation to be worse than more recent data, due to limited EHR use at this point in time (for example, chemotherapy prescriptions were not completely electronic until 2012), limiting rapid NLP/structured data extraction. Finally, RWD relies on source data entered by healthcare professionals being accurate when in reality data may sometimes be entered erroneously or not at all. Corroborating multiple data sources for each data category ensured each patient’s data was as complete and accurate as feasible.

A random selection of 60 of 2895 patients for key categories was prioritised, therefore accuracy values recorded are estimations. Due to the dataset size and resources available, it was not feasible to carry out a more extensive data validation and verification process, and therefore sampling was undertaken as a pragmatic approach. This is an issue that has been highlighted with RWD validation processes even by large commercial companies with much larger resources (2). Data was also constantly checked during the data curation process to ensure robustness.

### Implications and Future work

RT-HaND can now support internal and external single- or multi-modal original or validation research projects, and we invite collaborators, with our data available on request (https://github.com/GSTT-Radiotherapy-Physics/RT-HaND/tree/main). Extensive Standard Operating Procedure documentation is available on GitHub. RT-HaND can enable researchers and clinicians to derive reliable insights, from improving prognostic models to examining real-world treatment outcomes, ultimately facilitating data-driven improvements in care.

Work is ongoing to continuously improve the dataset, both by addressing any areas of missing data for the initial cohort, but prospectively updating data for the initial cohort and adding new patients using automated EDW queries to interrogate prospective data recorded in our centre’s new unified EHR system. This is a ‘live’ dataset that will undergo regular updates (and evaluation) and iterative improvement, therefore serving as a dynamic research resource. We hope that future NLP advancements may be incorporated into resolving current dataset gaps. We also hope to expand the horizons of our dataset too by soon incorporating biobank tissue data for further data aspects including genomics.

## Conclusions

RT-HaND-C represents one of the most extensive and robust clinical HNC real-world datasets available, comprising demographic, co-morbidity, diagnosis, treatment, laboratory and outcome data for a complete HNC oncology cohort using a combination of data curation strategies including EHR structured data extracts and AI. Our evaluation framework, showing RT-HaND-C to possess excellent accuracy, completeness and consistency of critical variables, can be followed as a template for other RWD quality assessments. RT-HaND-C is a ‘live’ dataset with ongoing improvements and prospective data additions with automated data pipelines.

Virtual linkage to RT-HaND-I ensures this dataset can underpin many future multi-modal research projects. We welcome collaboration to further harness RT-HaND-C’s potential to drive forward evidence-based improvements in HNC care.

## Data Availability

All data produced in the present study are available upon reasonable request to the authors.

https://github.com/GSTT-Radiotherapy-Physics/RT-HaND/tree/main/Documentation

