## Supplementary Tables for "RT-HaND-C: A Multi-Source, Validated Real-World Head and Neck Cancer Dataset for Research"

| **Concept** |
|---|
| Atrial fibrillation (disorder) |
| Block dissection of cervical lymph nodes |
| Carboplatin |
| Cetuximab |
| Chemotherapy |
| Chronic disease of respiratory system (disorder) |
| Chronic kidney disease (disorder) |
| Chronic liver disease (disorder) |
| Cisplatin |
| Combined chemotherapy and radiation therapy |
| Combined post-operative chemotherapy and radiotherapy |
| Complete therapeutic response (finding) |
| Curative - procedure intent (qualifier value) |
| Diabetes mellitus (disorder) |
| Docetaxel |
| Enteral feeding |
| Excision of mandible (procedure) |
| Exteriorization of trachea |
| Fluorouracil |
| Gastrostomy tube |
| Heart disease (disorder) |
| Human papillomavirus negative squamous cell carcinoma |
| Human papillomavirus positive squamous cell carcinoma |
| Hypertensive disorder, systemic arterial (disorder) |
| Induction chemotherapy |
| Laryngectomy |
| Malignant tumor of larynx |
| Malignant tumor of oropharynx |
| Maxillectomy (procedure) |
| Nasogastric tube, device |
| Nivolumab (substance) |
| No evidence of recurrence of cancer |
| Palliative - procedure intent (qualifier value) |
| Palliative course of radiotherapy (procedure) |
| Partial glossectomy (procedure) |

| |
|---|
| Perineural invasion by tumor present |
| Postoperative course of radiotherapy |
| Primary squamous cell carcinoma of base of tongue (disorder) |
| Primary squamous cell carcinoma of glottis (disorder) |
| Primary squamous cell carcinoma of palatine tonsil (disorder) |
| Primary squamous cell carcinoma of retromolar area (disorder) |
| Primary squamous cell carcinoma of soft palate (disorder) |
| Primary squamous cell carcinoma of uvula (disorder) |
| Radiation oncology AND/OR radiotherapy |
| Salvage procedure (qualifier value) |
| Squamous cell carcinoma |
| Squamous cell carcinoma of buccal mucosa (disorder) |
| Squamous cell carcinoma of larynx |
| Squamous cell carcinoma of lip (disorder) |
| Squamous cell carcinoma of tongue |
| Tonsillectomy (procedure) |

**Supplementary Table 1. SNOMED CT terms searched for the patient population using CogStack (validated previously during CogStack evaluation project (25)).**

| Gender | n | % |
|---|---|---|
| Female | 805 | 27.8% |
| Male | 2090 | 72.2% |

| Race Group | n | % |
|---|---|---|
| Asian Groups | 97 | 3.4% |
| Black Groups | 180 | 6.2% |
| White Groups | 1998 | 69.0% |
| Other Groups | 110 | 3.8% |
| Unknown | 510 | 17.6% |

| Age at Diagnosis | n | % |
|---|---|---|
| <40 | 96 | 3.3% |
| 40-49 | 298 | 10.3% |
| 50-59 | 753 | 26.0% |
| 60-69 | 898 | 31.0% |
| 70-79 | 576 | 19.9% |
| 80-89 | 237 | 8.2% |
| >90 | 37 | 1.3% |

| Co-morbidities | n | % |
|---|---|---|
| Hypertension | 1169 | 40.4% |
| Heart disease | 178 | 6.1% |
| Chronic resp disease | 588 | 20.3% |
| Chronic liver disease | 134 | 4.6% |
| Chronic kidney disease | 118 | 4.1% |
| Diabetes mellitus | 266 | 9.2% |
| Atrial fibrillation | 155 | 5.4% |
| NonHNCCancerDx | 275 | 9.5% |

| Year of Diagnosis | n | % |
|---|---|---|
| Pre-2010 | 40 | 1.4% |
| 2010 | 166 | 5.7% |
| 2011 | 205 | 7.1% |
| 2012 | 214 | 7.4% |
| 2013 | 201 | 6.9% |
| 2014 | 207 | 7.2% |
| 2015 | 215 | 7.4% |
| 2016 | 216 | 7.5% |
| 2017 | 214 | 7.4% |
| 2018 | 233 | 8.0% |
| 2019 | 222 | 7.7% |
| 2020 | 177 | 6.1% |
| 2021 | 227 | 7.8% |
| 2022 | 235 | 8.1% |
| 2023 (to 4th Oct) | 123 | 4.2% |

| HN Tumour Site | n | % |
|---|---|---|
| CUP | 99 | 3.4% |
| Hypopharynx | 152 | 5.3% |
| Lacrimal gland | 2 | 0.1% |
| Larynx | 568 | 19.6% |
| Multiple | 64 | 2.2% |
| Nasopharynx | 114 | 3.9% |
| Oral cavity | 641 | 22.1% |
| Oropharynx | 965 | 33.3% |
| Paranasal sinus/nasal cavity | 128 | 4.4% |
| Salivary gland | 162 | 5.6% |

| Status as of 4.10.23 | n | % |
|---|---|---|
| Alive | 1456 | 50.3% |
| Dead | 1439 | 49.7% |

| Histopathology | n | % |
|---|---|---|
| Acinar cell adenocarcinoma | 11 | 0.4% |
| Adenocarcinoma | 45 | 1.6% |
| Adenoid cystic carcinoma | 58 | 2.0% |
| Adenosquamous | 3 | 0.1% |
| Ameloblastic | 6 | 0.2% |
| Basal cell adenocarcinoma | 1 | 0.0% |
| Basaloid SCC | 3 | 0.1% |
| Carcinoma ex pleomorphic adenoma | 20 | 0.7% |
| Carcinoma NOS | 10 | 0.3% |
| Carcinosarcoma | 4 | 0.1% |
| Chondrosarcoma | 1 | 0.0% |
| In situ and dysplasia | 10 | 0.3% |
| Lymphoepithelial | 2 | 0.1% |

| | | |
|---|---|---|
| Melanoma | 3 | 0.1% |
| Merkel cell | 2 | 0.1% |
| Mucoepidermoid | 28 | 1.0% |
| Multiple - adenosquamous and SCC | 1 | 0.0% |
| Multiple - merkel and SCC | 1 | 0.0% |
| Multiple - SCC (sarcomatoid) and olfactory neuroblastoma | 1 | 0.0% |
| Myoepithelial | 7 | 0.2% |
| Neuroendocrine | 7 | 0.2% |
| Non-intestinal type adenocarcinoma | 1 | 0.0% |
| NPC | 82 | 2.8% |
| Olfactory neuroblastoma | 5 | 0.2% |
| Oncocytic carcinoma | 1 | 0.0% |
| Pleomorphic adenoma | 5 | 0.2% |
| Salivary duct carcinoma | 12 | 0.4% |
| SCC | 2518 | 87.0% |
| SCC (sarcomatoid) | 6 | 0.2% |
| SCC (spindle cell) | 4 | 0.1% |
| Small cell | 8 | 0.3% |
| SNUC | 8 | 0.3% |
| Transitional cell carcinoma | 1 | 0.0% |
| Undifferentiated | 20 | 0.7% |

| **cDisease Stage TNM7 at diagnosis** | **n** | **%** |
|---|---|---|
| 0 | 6 | 0.2% |
| Benign | 5 | 0.2% |
| I | 250 | 8.6% |
| II | 297 | 10.3% |
| III | 403 | 13.9% |
| IV | 1868 | 64.5% |
| Unknown | 66 | 2.3% |

| **Initial treatment received - summary** | **n** | **%** |
|---|---|---|
| Best supportive care | 117 | 4.0% |
| Definitive cetuximab + radiotherapy | 50 | 1.7% |
| Definitive chemoradiotherapy | 441 | 15.2% |
| Treatment or referred elsewhere | 43 | 1.5% |
| Definitive radiotherapy | 497 | 17.2% |
| Died before starting palliative treatment | 4 | 0.1% |

| | | |
|---|---|---|
| Induction chemotherapy + cetuximab + radiotherapy | 6 | 0.2% |
| Induction chemotherapy + chemoradiotherapy | 360 | 12.4% |
| Induction chemotherapy + definitive radiotherapy | 26 | 0.9% |
| Induction chemotherapy + palliative radiotherapy | 12 | 0.4% |
| Induction chemotherapy + surgery | 1 | 0.0% |
| Induction chemotherapy + surgery + post-operative radiotherapy | 4 | 0.1% |
| Induction chemotherapy then Cyberknife | 1 | 0.0% |
| Induction chemotherapy then died | 6 | 0.2% |
| Palliative radiotherapy | 435 | 15.0% |
| Palliative SACT | 61 | 2.1% |
| Surgery alone | 160 | 5.5% |
| Surgery + post-operative chemoradiotherapy | 169 | 5.8% |
| Surgery + post-operative radiotherapy | 497 | 17.2% |
| Surveillance | 5 | 0.2% |

| **HPV Status for oropharynx ca** | **n** | **%** |
|---|---|---|
| Negative | 207 | 21.5% |
| Positive | 597 | 62.0% |
| Unknown | 159 | 16.5% |

| **Induction chemo** | **n** | **%** |
|---|---|---|
| 0 | 2468 | 85.3% |
| 1 | 424 | 14.6% |
| Elsewhere | 3 | 0.1% |

| **Concomitant chemo** | **n** | **%** |
|---|---|---|
| 0 | 1842 | 63.6% |
| 1 | 1029 | 35.5% |
| Elsewhere | 24 | 0.8% |

| **Initial concomitant drug** | **n** | **%** |
|---|---|---|
| CARBOPLATIN | 169 | 16.0% |
| CETUXIMAB | 57 | 5.4% |
| Cis/etop | 2 | 0.2% |
| CISPLATIN | 808 | 76.7% |
| Olaparib trial | 3 | 0.3% |
| Unknown | 14 | 1.3% |

| **HNC RT** | **n** | **%** |
|---|---|---|
| 0 | 288 | 9.9% |
| 1 | 2557 | 88.3% |
| Elsewhere or historic | 50 | 1.7% |

| **HNC RT course 1 intent** | **n** | **%** |
|---|---|---|
| Radical | 2124 | 81.5% |
| Palliative | 483 | 18.5% |

**Supplementary Table 2. Dataset characteristics.**

| Demographic data category | Number assessed | Number correct | Accuracy |
|---|---|---|---|
| Date of Birth | 60 | 60 | 100.0% |
| Gender | 60 | 60 | 100.0% |
| Race/Ethnicity | 60 | 60 | 100.0% |
| Postcode | 60 | 59 | 98.3% |
| Address | 60 | 59 | 98.3% |
| Height | 60 | 60 | 100.0% |
| Weight (first recorded) | 60 | 60 | 100.0% |
| Date of Weight (first recorded) | 60 | 60 | 100.0% |
| Date of ECOG Performance Status (first recorded) | 60 | 60 | 100.0% |
| ECOG Performance Status (first recorded) | 60 | 60 | 100.0% |

**Supplementary Table 3. Accuracy estimation results for demographic data categories.**

| PMH data category | Number assessed | Number correct | Accuracy | PPV | NPV |
|---|---|---|---|---|---|
| Hypertension | 60 | 58 | 96.7% | 0.952 | 0.974 |
| Heart disease | 60 | 58 | 96.7% | Incalculable | 0.967 |
| Chronic respiratory disease | 60 | 60 | 100.0% | 1.000 | 1.000 |
| Chronic liver disease | 60 | 58 | 96.7% | 1.000 | 0.965 |
| Chronic kidney disease | 60 | 59 | 98.3% | 1.000 | 0.983 |
| Diabetes mellitus | 60 | 58 | 96.7% | 1.000 | 0.966 |
| Atrial fibrillation | 60 | 60 | 100.0% | 1.000 | 1.000 |
| Non-HNC Diagnosis 1 | 60 | 58 | 96.7% | 1.000 | 0.963 |
| Non-HNC Diagnosis 2 | 60 | 60 | 100.0% | Incalculable | 1.000 |

**Supplementary Table 4. Accuracy, PPV and NPV estimation results for co-morbidity data categories.**

| Dx data category | Number assessed | Number correct | Accuracy | PPV | NPV |
|---|---|---|---|---|---|
| Diagnosis Date | 60 | 60 | 100.0% | | |
| Oncology First Visit Date | 60 | 60 | 100.0% | | |
| Disease Major Site | 60 | 60 | 100.0% | | |
| Multiple HNC Sites | 60 | 60 | 100.0% | | |
| Disease Subsite | 25 | 25 | 100.0% | | |
| Disease Laterality | 29 | 27 | 93.1% | | |
| cTNM7 – T stage | 60 | 58 | 96.7% | | |
| cTNM7 – N stage | 60 | 57 | 95.0% | | |
| cTNM7 – M stage | 60 | 58 | 96.7% | | |
| cTNM7 – Overall Disease Stage | 60 | 57 | 95.0% | | |
| Lung metastases | 48 | 48 | 100.0% | 1 | 1 |
| Histopathology | 60 | 60 | 100.0% | | |
| Tumour Grade | 6 | 6 | 100.0% | | |
| HPV Status | 60 | 60 | 100.0% | 1 | 1 |
| EBV Status | 60 | 60 | 100.0% | 1 | 1 |
| Perineural invasion present | 20 | 17 | 85.0% | 0.5 | 1 |
| Surgical margin involvement | 5 | 5 | 100.0% | | |
| Extracapsular extension of nodal tumour | 5 | 5 | 100.0% | | |
| Lymphovascular invasion | 5 | 5 | 100.0% | | |
| PDL1 score (TPS) | 60 | 60 | 100.0% | | |
| PDL1 (TPS category) | 60 | 60 | 100.0% | | |
| PDL1 score (CPS) | 60 | 60 | 100.0% | | |
| PDL1 (CPS category) | 60 | 60 | 100.0% | | |
| Initial treatment received | 60 | 58 | 96.7% | | |

**Supplementary Table 5. Accuracy estimation results for disease characteristic and first treatment data categories. Data categories where less than 60 cases assessed reflects cases within the sample where data missing in dataset currently.**

| **Chemo data category** | **Number assessed** | **Number correct** | **Accuracy** | **PPV** | **NPV** |
|---|---|---|---|---|---|
| Neoadjuvant chemotherapy given | 60 | 60 | 100.0% | 1 | 1 |
| Neoadjuvant chemotherapy regimen 1 | 60 | 59 | 98.3% | | |
| Neoadjuvant chemotherapy cycle 1 date | 60 | 59 | 98.3% | | |
| Neoadjuvant cycles | 60 | 59 | 98.3% | | |
| Neoadjuvant cisplatin cycles | 60 | 59 | 98.3% | | |
| Neoadjuvant cisplatin total dose | 60 | 59 | 98.3% | | |
| Neoadjuvant carboplatin cycles | 60 | 59 | 98.3% | | |
| Neoadjuvant carboplatin total dose | 60 | 60 | 100.0% | | |
| Neoadjuvant 5FU cycles | 60 | 59 | 98.3% | | |
| Neoadjuvant 5FU total dose | 60 | 59 | 98.3% | | |
| Neoadjuvant Docetaxel cycles | 60 | 60 | 100.0% | | |
| Neoadjuvant Docetaxel total dose | 60 | 60 | 100.0% | | |
| Neoadjuvant Gemcitabine cycles | 60 | 60 | 100.0% | | |
| Neoadjuvant Gemcitabine total dose | 60 | 60 | 100.0% | | |
| Neoadjuvant chemotherapy regimen 2 | 60 | 60 | 100.0% | | |
| Concomitant chemotherapy given | 60 | 60 | 100.0% | 1 | 1 |
| Concomitant chemotherapy Drug 1 | 60 | 60 | 100.0% | | |
| Concomitant chemotherapy cycle 1 date | 60 | 60 | 100.0% | | |
| Concomitant chemotherapy Drug 1 total dose | 60 | 60 | 100.0% | | |
| Concomitant chemotherapy Drug 1 cycles | 60 | 60 | 100.0% | | |
| Concomitant chemotherapy Drug 1 dosing regimen | 60 | 60 | 100.0% | | |
| Concomitant chemotherapy Drug 2 | 60 | 60 | 100.0% | | |
| Concomitant chemotherapy Drug 2 total dose | 60 | 60 | 100.0% | | |
| Concomitant chemotherapy Drug 2 cycles | 60 | 60 | 100.0% | | |
| Concomitant chemotherapy Drug 2 dosing regimen | 60 | 60 | 100.0% | | |
| Palliative SACT given | 60 | 60 | 100.0% | 1 | 1 |
| 1$^{st}$ line palliative SACT regime | 60 | 60 | 100.0% | | |
| 1$^{st}$ line palliative SACT cycle 1 date | 60 | 60 | 100.0% | | |
| 1$^{st}$ line palliative SACT cycles | 60 | 60 | 100.0% | | |

| | | | |
|---|---|---|---|
| 2nd line palliative SACT regime | 60 | 60 | 100.0% |
| 2nd line palliative SACT cycle 1 date | 60 | 60 | 100.0% |
| 2nd line palliative SACT cycles | 60 | 60 | 100.0% |
| 3rd line palliative SACT regime | 60 | 60 | 100.0% |
| 3rd line palliative SACT cycle 1 date | 60 | 60 | 100.0% |
| 3rd line palliative SACT cycles | 60 | 60 | 100.0% |
| 4th line palliative SACT regime | 60 | 60 | 100.0% |
| 4th line palliative SACT cycle 1 date | 60 | 60 | 100.0% |
| 4th line palliative SACT cycles | 60 | 60 | 100.0% |
| 5th line palliative SACT regime | 60 | 60 | 100.0% |
| 5th line palliative SACT cycle 1 date | 60 | 60 | 100.0% |
| 5th line palliative SACT cycles | 60 | 60 | 100.0% |

**Supplementary Table 6. Accuracy estimation results for chemotherapy data categories.**

| RT data category | Assessed | Correct | Accuracy | PPV | NPV |
|---|---|---|---|---|---|
| HNC RT given | 60 | 60 | 100.0% | 1 | 1 |
| HNCRT course 1 site | 60 | 60 | 100.0% | 1 | 1 |
| HNC RT course 1 intent | 60 | 60 | 100.0% | | |
| HNC RT course 1 dose per Fr | 60 | 59 | 98.3% | | |
| HNC RT course 1 recorded total dose | 60 | 59 | 98.3% | | |
| HNC RT course 1 recorded fractions | 60 | 58 | 96.7% | | |
| HNC RT course 1 initially intended total dose | 60 | 59 | 98.3% | | |
| HNC RT course 1 initially intended fractions | 60 | 59 | 98.3% | | |
| HNC RT course 1 start date | 60 | 60 | 100.0% | | |
| HNC RT course 1 end date | 60 | 60 | 100.0% | | |
| HNC RT course 1 elapsed time | 60 | 60 | 100.0% | | |
| HNC RT course 1 replans | 60 | 59 | 98.3% | 1 | 0.982 |
| HNC RT course 1 early termination | 60 | 59 | 98.3% | 1 | 0.982 |
| HNC RT course 1 early termination reason | 60 | 59 | 98.3% | | |
| HNCRT course 2 site | 60 | 60 | 100.0% | | |
| HNC RT course 2 intent | 60 | 60 | 100.0% | | |
| HNC RT course 2 dose per Fr | 60 | 60 | 100.0% | | |
| HNC RT course 2 total dose | 60 | 60 | 100.0% | | |

| | | | |
|---|---|---|---|
| HNC RT course 2 fractions | 60 | 60 | 100.0% |
| Non-HNC RT | 60 | 60 | 100.0% |

**Supplementary Table 7. Accuracy estimation results for RT data categories.**

| Surgical data category | Number assessed | Number correct | Accuracy | PPV | NPV |
|---|---|---|---|---|---|
| Laryngectomy | 60 | 59 | 98.3% | 1.000 | 0.982 |
| Laryngectomy Procedure Description | 60 | 59 | 98.3% | | |
| Laryngectomy Procedure Date | 60 | 59 | 98.3% | | |
| Neck dissection | 60 | 57 | 95.0% | 0.952 | 0.949 |
| Neck dissection Procedure Description | 60 | 57 | 95.0% | | |
| Neck dissection Procedure Date | 60 | 57 | 95.0% | | |
| Tracheostomy | 60 | 59 | 98.3% | 1.000 | 0.978 |
| Tracheostomy Procedure description | 60 | 59 | 98.3% | | |
| Tracheostomy First Procedure Date | 60 | 59 | 98.3% | | |
| Mandible excision | 60 | 59 | 98.3% | 1.000 | 0.982 |
| Mandible excision Procedure Description | 60 | 59 | 98.3% | | |
| Mandible excision Procedure Date | 60 | 59 | 98.3% | | |
| Maxillectomy | 60 | 60 | 100.0% | 1.000 | 1.000 |
| Maxillectomy Procedure Description | 60 | 60 | 100.0% | | |
| Maxillectomy Procedure Date | 60 | 60 | 100.0% | | |
| Total Glossectomy | 60 | 60 | 100.0% | 1.000 | 1.000 |
| Total Glossectomy Procedure Description | 60 | 60 | 100.0% | | |
| Total Glossectomy Procedure Date | 60 | 60 | 100.0% | | |

| | | | | | |
|---|---|---|---|---|---|
| Partial Glossectomy | 60 | 60 | 100.0% | 1.000 | 1.000 |
| Partial Glossectomy Procedure Description | 60 | 60 | 100.0% | | |
| Partial Glossectomy Procedure Date | 60 | 60 | 100.0% | | |
| Oral cavity surgery | 60 | 60 | 100.0% | 1.000 | 1.000 |
| Oral cavity surgery Procedure Description | 60 | 60 | 100.0% | | |
| Oral cavity surgery Procedure Date | 60 | 60 | 100.0% | | |
| Tonsillectomy | 60 | 60 | 100.0% | 1.000 | 0.982 |
| Tonsillectomy Procedure Description | 60 | 60 | 100.0% | | |
| Tonsillectomy Procedure Date | 60 | 60 | 100.0% | | |
| Pharyngectomy | 60 | 60 | 100.0% | 1.000 | 1.000 |
| Pharyngectomy Procedure Description | 60 | 60 | 100.0% | | |
| Pharyngectomy Procedure Date | 60 | 60 | 100.0% | | |
| Salivary gland surgery | 60 | 59 | 98.3% | 1.000 | 0.982 |
| Salivary gland surgery procedure description | 60 | 59 | 98.3% | | |
| Salivary gland surgery First Procedure Date | 60 | 59 | 98.3% | | |
| Nasal cavity sinus surgery | 60 | 59 | 98.3% | 1.000 | 1.000 |
| Nasal cavity sinus surgery Procedure Description | 60 | 60 | 100.0% | | |
| Nasal cavity sinus surgery Procedure Date | 60 | 60 | 100.0% | | |
| Orbital Exenteration | 60 | 60 | 100.0% | 1.000 | 1.000 |
| Orbital Exenteration Procedure Description | 60 | 60 | 100.0% | | |
| Orbital Exenteration Procedure Date | 60 | 60 | 100.0% | | |
| Salvage procedure CogStack | 50 | 49 | 98.0% | 1.000 | 0.981 |
| Electrochemotherapy | 60 | 60 | 100.0% | | |

| | | | | | |
|---|---|---|---|---|---|
| Electrochemotherapy procedure description | 60 | 60 | 100.0% | | |
| Electrochemotherapy first Procedure Date | 60 | 60 | 100.0% | | |
| Gastrostomy | 60 | 58 | 96.7% | 0.955 | 0.974 |
| Gastrostomy procedure description | 60 | 58 | 96.7% | | |
| Gastrostomy date | 60 | 58 | 96.7% | | |
| Gastrostomy removal | 14 | 14 | 100.0% | 1.000 | 1.000 |
| Gastrostomy removal Procedure Description | 14 | 14 | 100.0% | | |
| Gastrostomy removal First Procedure Date | 14 | 14 | 100.0% | | |

**Supplementary Table 8. Accuracy estimation results for Surgical data categories. Data categories where less than 60 cases assessed reflects cases within the sample where data missing in dataset currently.**

| Outcomes data point | Number assessed | Number correct | Accuracy |
|---|---|---|---|
| Last Follow-up Date | 60 | 59 | 98.3% |
| Deceased | 60 | 60 | 100.0% |
| Death Date | 60 | 60 | 100.0% |
| Cause of death | 3 | 3 | 100% |
| Cause of death other | 3 | 3 | 100% |
| Complete Response (CogStack ‘complete response’ + radical RT) | 9 | 9 | 100% |
| 3 month RT response | 30 | 30 | 100% |
| 3 month imaging modality | 18 | 18 | 100% |
| 3 month imaging date | 24 | 24 | 100% |
| 6 month RT response | 27 | 27 | 100% |
| 6 month imaging modality | 20 | 20 | 100% |
| 6 month imaging date | 22 | 22 | 100% |
| Biopsy post-RT | 0 | 0 | n/a |
| Biopsy date | 0 | 0 | n/a |
| Salvage neck dissection | 13 | 13 | 100% |
| Salvage neck dissection date | 13 | 13 | 100% |
| Failure | 18 | 17 | 94.4% |
| Failure date | 15 | 15 | 100% |
| Failure site summary | 15 | 15 | 100% |
| Primary failure site details | 13 | 13 | 100% |
| Primary recurrence date | 13 | 13 | 100% |
| Primary recurrence Treatment | 13 | 13 | 100% |
| Primary recurrence intent | 13 | 13 | 100% |
| Nodal or locoregional failure site | 14 | 14 | 100% |
| Nodal or locoregional non-primary recurrence date | 14 | 14 | 100% |
| Nodal recurrence Treatment | 13 | 13 | 100% |
| Nodal recurrence intent | 13 | 13 | 100% |
| Metastatic site | 13 | 13 | 100% |
| Metastatic recurrence date | 14 | 14 | 100% |
| Metastatic recurrence Treatment | 14 | 14 | 100% |
| Metastatic recurrence intent | 14 | 14 | 100% |
| Recurrence after RT | 19 | 19 | 100% |
| Time to failure after RT | 19 | 19 | 100% |
| In-field recurrence (high dose region) | 17 | 17 | 100% |
| In-field recurrence (prophylactic dose region) | 17 | 17 | 100% |

**Supplementary Table 9. Accuracy estimation results for outcome data categories. Data categories where less than 60 cases assessed reflects cases within the sample where data missing in dataset currently.**

| | Spearman Correlation Coefficient versus Max acute period %Weight change | Lower 95% CI | Upper 95% CI | Sig. (2-tailed) | n |
|---|---|---|---|---|---|
| %Weight change 6 week post-RT | 0.719 | 0.676 | 0.757 | <.001 | 588 |
| %Weight change 3-months post-RT | 0.577 | 0.519 | 0.63 | <.001 | 601 |
| %Weight change 6-months post-RT | 0.431 | 0.35 | 0.505 | <.001 | 452 |
| %Weight change 1-year post-RT | 0.323 | 0.234 | 0.406 | <.001 | 439 |
| %Weight change 2-year post-RT | 0.276 | 0.168 | 0.377 | <.001 | 321 |
| %Weight change 3-year post-RT | 0.29 | 0.156 | 0.414 | <.001 | 206 |
| %Weight change 4-year post-RT | 0.13 | -0.029 | 0.282 | 0.098 | 163 |
| %Weight change 5-year post-RT | 0.327 | 0.159 | 0.476 | <.001 | 130 |

**Supplementary Table 10. Spearman's rank correlation between maximum acute period percentage weight loss from baseline compared to percentage weight change from baseline later timepoints. Significant at adjusted significance value of 0.00625 (Bonferroni correction for 8 comparisons, green highlighting denotes significance).**
